# Demographic analysis of mortality changes associated with the COVID-19 pandemic in Japan, 2020–2022

**DOI:** 10.1101/2025.04.13.25325456

**Authors:** Yuta Okada, Hiroshi Nishiura

## Abstract

**Objectives:** To evaluate the mortality impact of the COVID-19 pandemic by age and prefecture in Japan.

**Study design:** Retrospective demographic analysis.

**Methods:** A hierarchical time-varying regression model was applied to prefectural life table data in Japan during 2000–2019. Pre-pandemic baseline mortality rates and death counts during 2020–2022 were projected by this model to evaluate the mortality impact of the COVID-19 pandemic on mortality during 2020–2022.

**Results:** Nationally, the observed mortality rates were higher than projected rates among those aged 10–29 years and lower for ages 1–9 and >80 years in 2020. During 2021 and 2022, mortality rates consistently increased for most age groups, with substantial regional heterogeneity in those aged <14 years. Consistently, the neutral gaps between the observed and projected life expectancy across prefectures in 2020 turned negative in most prefectures during 2022. In 2020, the negative gap in life expectancy ranged from a substantial shortening of −1.41 years (95% prediction interval [PI]: −1.69, −1.12) in Okinawa to a slight shortening of −0.23 years (95% PI: −0.63, 0.17) in Tottori. Excess deaths per population also grew consistently in all prefectures during 2020–2022; Miyazaki yielded the largest estimate at 186.2 (95% PI: 158.6, 214.0) per 100,000 population.

**Conclusions:** Our framework highlights the usefulness of life table data for evaluating the mortality impact caused by an event such as the pandemic. Our estimates, adjusted by pre-pandemic demographic trends, revealed the massive mortality impact of the COVID-19 pandemic across a wide age range.

## Introduction

COVID-19 has caused substantial morbidity and mortality worldwide since its emergence in Wuhan, China in November 2019. In addition to documented COVID-19 deaths, substantial mortality directly or indirectly associated with COVID-19 has stemmed from various mechanisms, including cardiovascular complications or limited health care access.^1–4^ At the global level, an estimated 14.9–15.9 million excess deaths occurred up to the end of 2021^5–7^, with excess deaths consistently reported even after 2022. ^8,9^ Trends in life expectancy estimates are also in line with those suggested by the excess mortality estimates, elucidating a substantial impact on mortality during the early phases of the COVID-19 pandemic in many countries. ^6,10–13^

Globally, evaluation of the total mortality impact owing to the COVID-19 pandemic has been carried out using excess mortality estimates. This approach is widely accepted for quantifying the mortality impact of epidemics, such as seasonal influenza, which is not fully observable by relying on indicator-based surveillance. Technically, excess mortality, expressed either as an absolute number or rate per population, is typically estimated as the difference between the observed and predicted mortality, or the “baseline” mortality projected by a statistical model fitted to the historical baseline.^5,6,14–20^ The definition of “baseline” mortality is a practical issue in its application to COVID-19. The baseline is updated according to the latest available data in widely used approaches for the calculation of excess mortality; thus, in the case of COVID-19, the baseline spans an extended period. This leads to difficulty in interpreting the estimated excess deaths as the genuine impact of mortality owing to the pandemic. One such example is the weekly excess mortality estimates in Japan provided in the “Excess and Exiguous Deaths Dashboard in Japan” (URL: https://exdeaths-japan.org/en/; hereinafter, ExDeath Dashboard).^9^ In that data source, the excess mortality at national and prefectural levels is calculated using the Farrington algorithm, which updates the baseline to the latest 5-year period of available weekly data.^9,18^

As for the estimation of excess mortality, another practical issue in conventional approaches is that these estimates do not provide results that are age-standardized or age-stratified. This can potentially lead to inappropriate interpretation of the mortality impact caused by external factors including COVID-19, especially when the underlying demographic change is not negligible. One measure to circumvent this issue is to deploy demographic approaches based on life table data to capture the temporal change in the age-wise mortality structure as well as to forecast mortality.^21–24^ Compared with conventional excess mortality estimates on a weekly basis, an apparent limitation of a life table-based approach is that life table data are usually only available on a yearly basis, with a certain time delay in reporting. However, an obvious strength of a life table-based approach is that key backbones in the age-mortality correlation and the underlying demographic trends can be captured, which may enable technically more sound projections of pre-pandemic trends in mortality beyond 2020, compared with existing approaches.

Given the above background, in the present study, we aimed to establish a novel framework to estimate post-pandemic changes in mortality by age group at national and subnational levels using longitudinal life table data provided in the Japanese Mortality Database.^25^ Using life table data during 2000–2019, we applied a time-varying regression model with a hierarchical structure that captured the pre-pandemic trend in mortality schedules by age group. The mortality during 2020–2022 projected by this model revealed the changes in mortality and life expectancy attributable to the COVID-19 pandemic during 2020–2022 at national and prefectural levels.

## Methods

### Epidemiological data

We retrieved national and prefectural life table data in Japan from 2020 to 2022 from The Japanese Mortality Database. ^25^ We recalculated these life tables in order to handle those over 100 in a single age group. Death counts in each age group (not integers) were rounded to the closest integer values for subsequent analysis.

We retrieved data on mortality caused by the 2011 Tohoku earthquake from the vital statistics of 2011 published by the Ministry of Health, Labour and Welfare. ^26^ For subsequent statistical inference, we basically subtracted disaster-associated death counts owing to the Tohoku earthquake from age groups >5 years for Iwate, Miyagi, and Fukushima prefectures. In the Tohoku earthquake data, the youngest age group was categorized as 0–4 years, which does not match the classification in the life tables (ages 0 and 1–4 as separate groups); we therefore accounted for the disaster-related mortality in the statistical model.

We also retrieved estimated weekly excess or exiguous death estimates from the ExDeath Dashboard^9^ during 2020–2022 for comparison with our excess death estimates.

### Decomposition of the national-level mortality table during 2000–2019

We used the age-stratified mortality rates from national life tables during 2000–2019 to compose a mortality schedule matrix **M**. Singular value decomposition of **M** is a widely used method in demography to extract key characteristics in mortality schedules. The result of decomposition can be written in the form

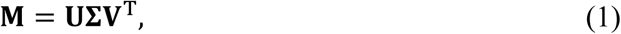

where **U** is a time-dependent component, 𝚺 generates singular values, and **V** contains column vectors that represent key structures in mortality along age. We applied singular value decomposition to the national longitudinal mortality schedule matrix, rather than to the mortality schedule matrix obtained by combining data from all regions, as in Alexander et al.^24^

### Hierarchical time-varying regression model

We built a statistical model to capture national-level mortality trends while also allowing variability by prefecture, similar to that in Alexander et al., with some modifications.^24^ Briefly, we modeled the mortality rate in age group *x* and in prefecture *a* in year *t* as a linear combination of 𝑌*_x_*^(1)^, 𝑌*_x_*^(2)^, and 𝑌*_x_*^(3)^, which are the three leading column vectors of **V** described above:

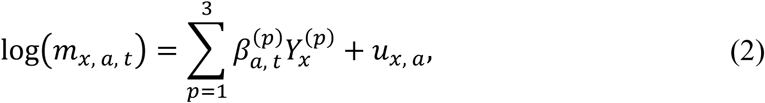

where 𝛽*_a,t_*^(𝑝)^ (𝑝 = 1, 2, 3) are the time-varying coefficients in year *t*. For details on the shapes and reason for choosing 𝑌*_x_*^(1)^, 𝑌*_x_*^(2)^, and 𝑌*_x_*^(3)^, see the Supplementary Methods 3. The final term 𝑢_𝑥, 𝑎_ in equation (2) is the random effects that were assumed to be time-invariant in our model. We also assumed that 𝛽*_a,t_*^(𝑝)^ is modeled as the sum of the national averages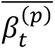 and random effects 𝜓_𝛽|a,t_^(𝑝)^ in year *t*; see the Supplementary Methods 1 for details.

A main part of the likelihoods considered in the present study was based on the observation process of death counts in each prefecture and in each age group. We assumed that the observed mortality rate in age group *x*, prefecture *a*, in year *t* follows a Poisson distribution. Additionally, the likelihood for total death counts across all age groups in each prefecture was also taken into consideration.^27^ See the Supplementary Methods 1 on how the effect of the 2011 earthquake is accounted for with respect to *m*′_𝑥,𝑎,𝑡_.

### Statistical inference of temporal changes in mortality

The statistical model for the temporal changes in 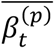 was dealt with as time-dependent and was assumed as a second-order random walk:

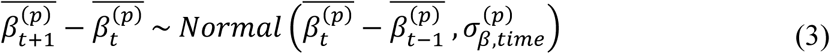

We used the Markov chain Monte Carlo method to estimate unknown parameters.

On the basis of the estimates of 𝛽*_a,t_*^(𝑝)^ from the 2000–2019 data, we projected 𝛽*_a,t_*^(𝑝)^ up to 2022. Therefore, we assumed the following: (i) the change rate of 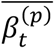 from 2018 to 2019 was maintained up to 2022, (ii) the prefectural random effects 𝜓_𝛽|𝑎,2022_^(𝑝)^, 𝜓_𝛽|𝑎,2021_^(𝑝)^, and 𝜓_𝛽|𝑎,2020_^(𝑝)^ are equal to 𝜓_𝛽|𝑎,2019_^(𝑝)^; consequently, we obtained the projected mortality rates at the national level, 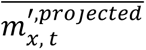, and at prefectural levels, 𝑚_𝑥, 𝑎, 𝑡_^′𝑝𝑟𝑜𝑗𝑒𝑐𝑡𝑒𝑑^, up to 2022. Excess or exiguous deaths were computed as the gap between projection-based prediction and observations of death counts during 2020–22. The gaps between observed and projected life expectancy at birth, 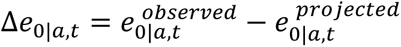, were also evaluated by calculating 𝑒_0|𝑎,𝑡_^𝑝𝑟𝑜𝑗𝑒𝑐𝑡𝑒𝑑^ using predicted values of 𝑚_𝑥, 𝑎, 𝑡_^′𝑝𝑟𝑜𝑗𝑒𝑐𝑡𝑒𝑑^ (𝑡 = 2020-2022). The calculation of life expectancy was based on standard methods to calculate period life tables^28^. See the Supplementary Methods 1-2 for further technical details.

### Statistical analysis software

All statistical and numerical analyses in the present study were performed using R version 4.2.2 (The R Project for Statistical Computing, Vienna, Austria) and CmdStan version 2.33.0.^29,30^

## Results

Changes in the national mortality rates by age group during 2019–2022 are shown in Figure 1. The mortality curves during 2019–2022 were not substantially different and were lower overall across all age groups, compared with 2000 or 2010 (Figure 1A). The difference in mortality curves during 2020-2022 compared with that of 2019 is shown in Figure 1B. During 2020–2022, the overall mortality rates among those younger than 10 years old were lower than those in 2019, which contrasts with the higher mortality rates during the same period for individuals in their 10s and 20s. In age groups >30 years, 2022 is outstanding in that mortality rates among all age groups >30 years were higher than those in 2019.

**Figure 1.**
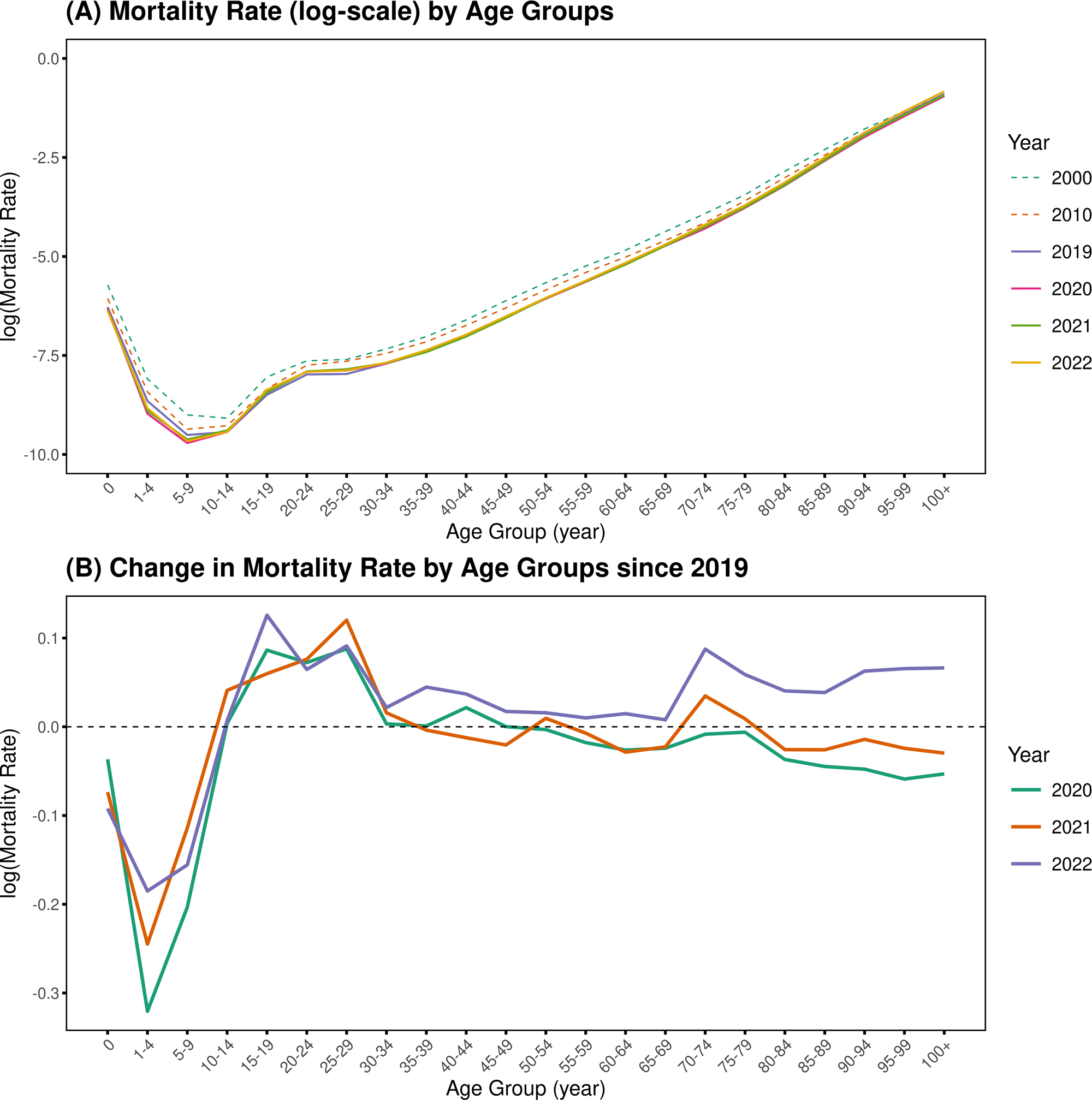
Changes in mortality rates by age group during 2000–2022. Panel (A) shows mortality schedule curves in 2020–2022 in comparison with those in 2000, 2010, and 2019. Panel (B) provides a closer look at the change in mortality rates by age group in 2020–2022 compared with that in 2019.

The difference between the observed and projected mortality rates in 2020– 2022 is shown in Figure 2. The overall trend seen in the results from 2020 (panel A) is substantial exiguous mortality in those aged 1–9 and ≥80 years and substantial excess mortality in those aged 15–59 and 65–74 years. This substantial excess mortality rate became greater in magnitude during 2021 and 2022, and the age range with excess mortality rate widened up to those aged 75–79 years. The exiguous mortality rate among individuals aged 1–9 years disappeared in 2021 and 2022 whereas that among those aged ≥80 years was still observable among individuals aged ≥95 years.

**Figure 2.**
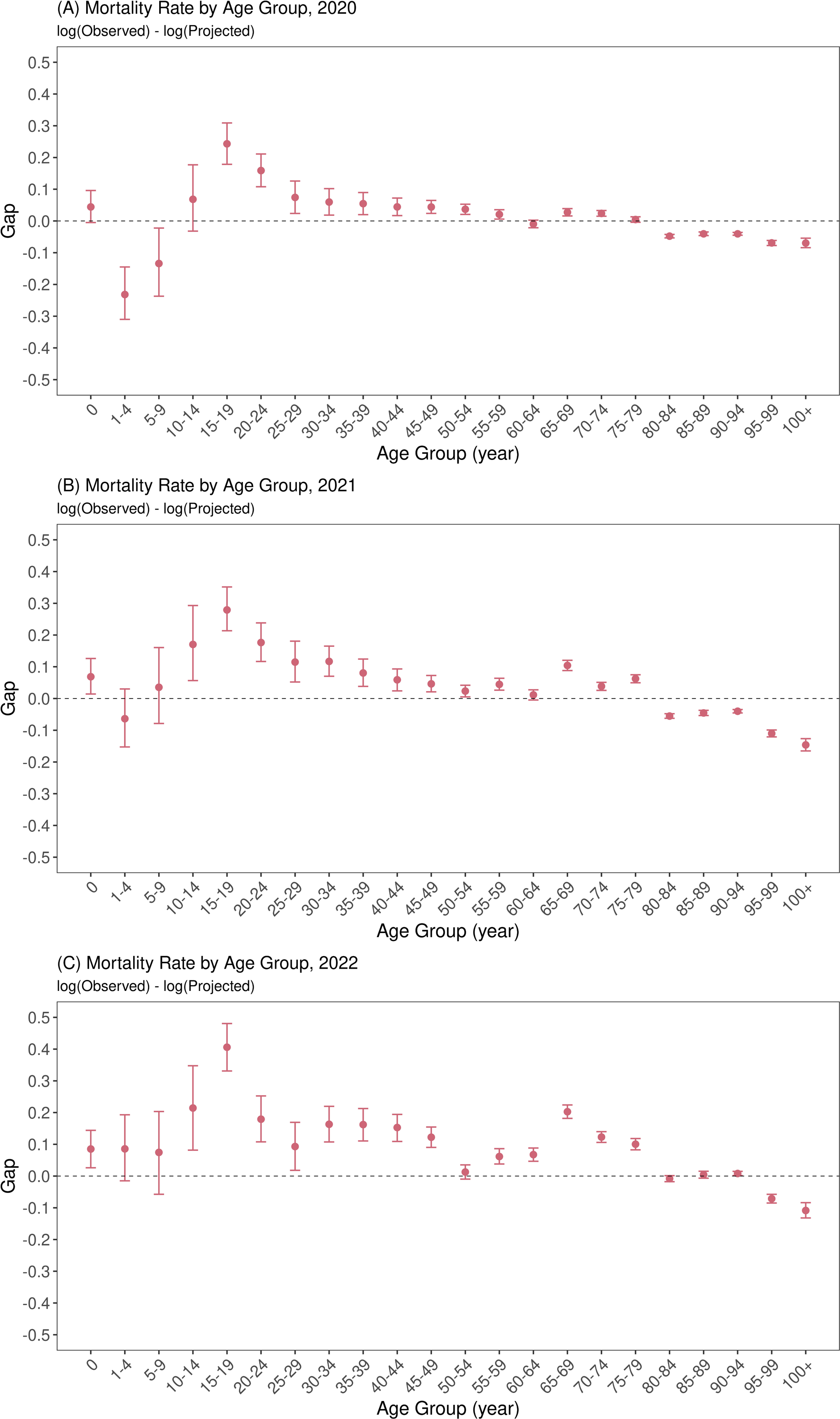
Gaps between observed and projected mortality rates by age group, 2020–2022. The red points with error bars show the median and 95% prediction intervals of the gaps between observed and projected (log-scaled) mortality rates in (A) 2020, (B) 2021, and (C) 2022.

The prefectural version of the values for the calculated gap between observed and projected mortality rates during 2020–2022 is shown in Figure 3. The overall tendency during 2020–2022 is a gradual increase in these gaps among those aged ≥30 years, where most age groups experienced consistent increases in mortality. Conversely, greater heterogeneity among prefectures was observed in those younger than 30 years old for the entire period, but the overall trend showed an increase in excess mortality, even in those younger age groups.

**Figure 3.**
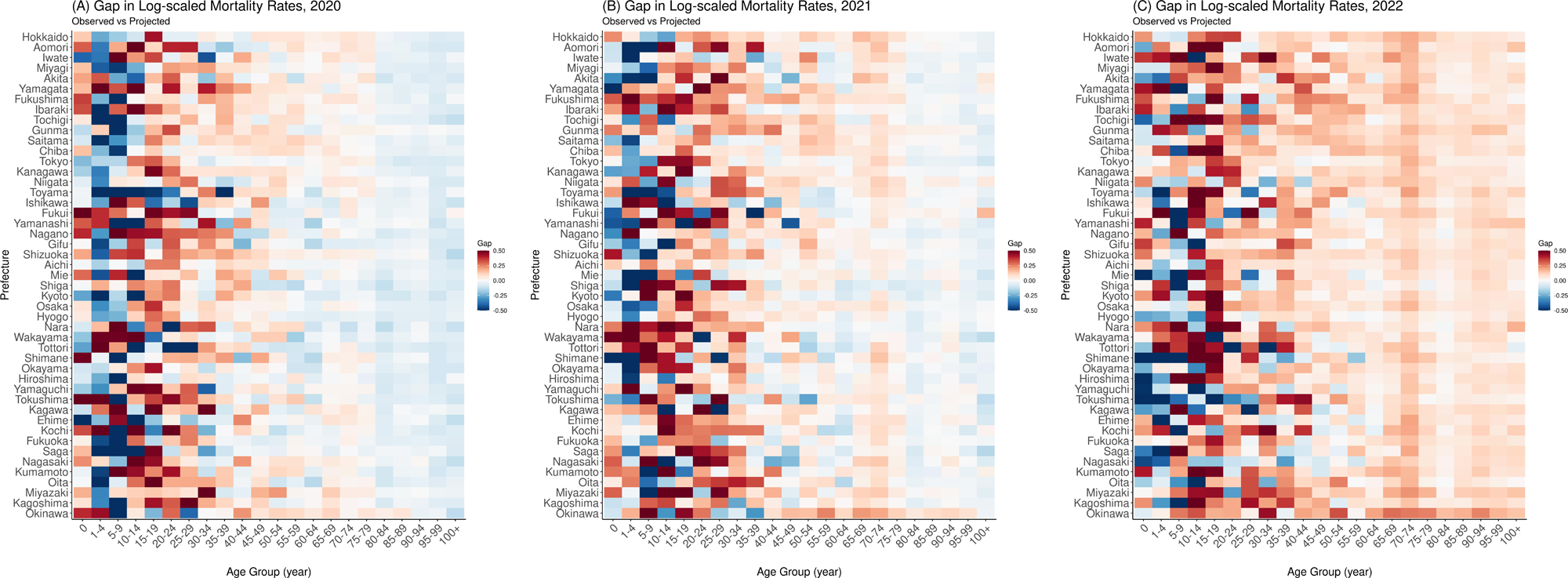
Heatmaps of the gap between observed and projected mortality rates by prefecture in 2020–2022. The cells represent gaps between the observed and the median of projected (log-scaled) mortality rates in (A) 2020, (B) 2021, and (C) 2022 for all prefectures. Cells with colors closer to dark blue show mortality rates that were lower than expected (projected) whereas those closer to solid red indicate mortality rates that were higher than expected.

A real-time update of the gap between observed and projected life expectancy at birth is depicted in Figure 4, showing how long the life expectancy was shortened in each pandemic year. In 2020, the life expectancy gaps were neutral to positive in all except a few prefectures. The finding in 2021 differed in that these gaps became mostly neutral to negative, and only the gap in Iwate was clearly in the positive range. The negative tendency grew greater in 2022 when these gaps were clearly negative in all prefectures except Tottori, where the gap was −0.235 (95% prediction interval [PI]: −0.631, 0.168). The greatest negative gap was −1.41 years (95% PI: −1.69, −1.12) in Okinawa.

**Figure 4.**
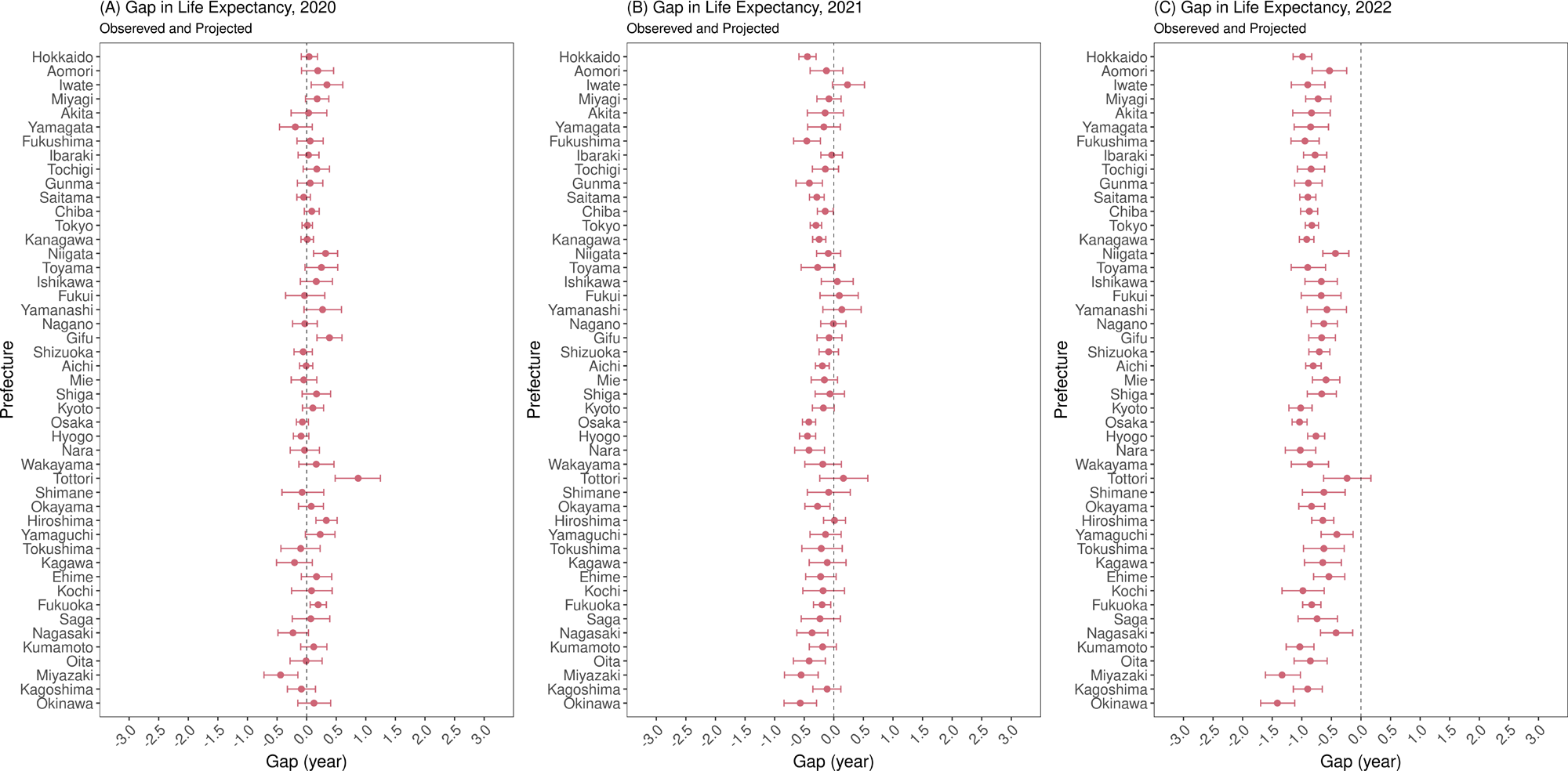
Gaps between life expectancy at birth calculated from observed and projected mortality rates in 2020–2022. Gaps between observed and projected life expectancy at birth are shown in (A) 2020, (B) 2021, and (C) 2022 for all prefectures. In each panel and for each prefecture, red points and error bars show the median and 95% prediction interval, respectively.

Figure 5 shows the mortality gap between observed and projected deaths per 100,000 population in all prefectures, in comparison with the yearly aggregated mean predicted gaps provided in ExDeath dashboard.^9^ Our estimates showed neutral to exiguous deaths in most prefectures in 2020 (Figure 5A). However, neutral to excess deaths were seen in 2021, and this trend grew worse in 2022 when all prefectures experienced substantial excess deaths. The worst prefecture in terms of excess deaths was Miyazaki with 186.2 (95% PI: 158.6, 214.0) excess deaths per 100,000 population; this was followed by Okinawa (146.9, 95% PI: 127.5, 166.1) and Toyama (145.5, 95% PI: 117.2, 174.0). Comparison with the data from ExDeath Dashboard in 2022 revealed some discrepancy with our estimates, especially in highly populated prefectures including Tokyo, Osaka, and their neighboring prefectures.

**Figure 5.**
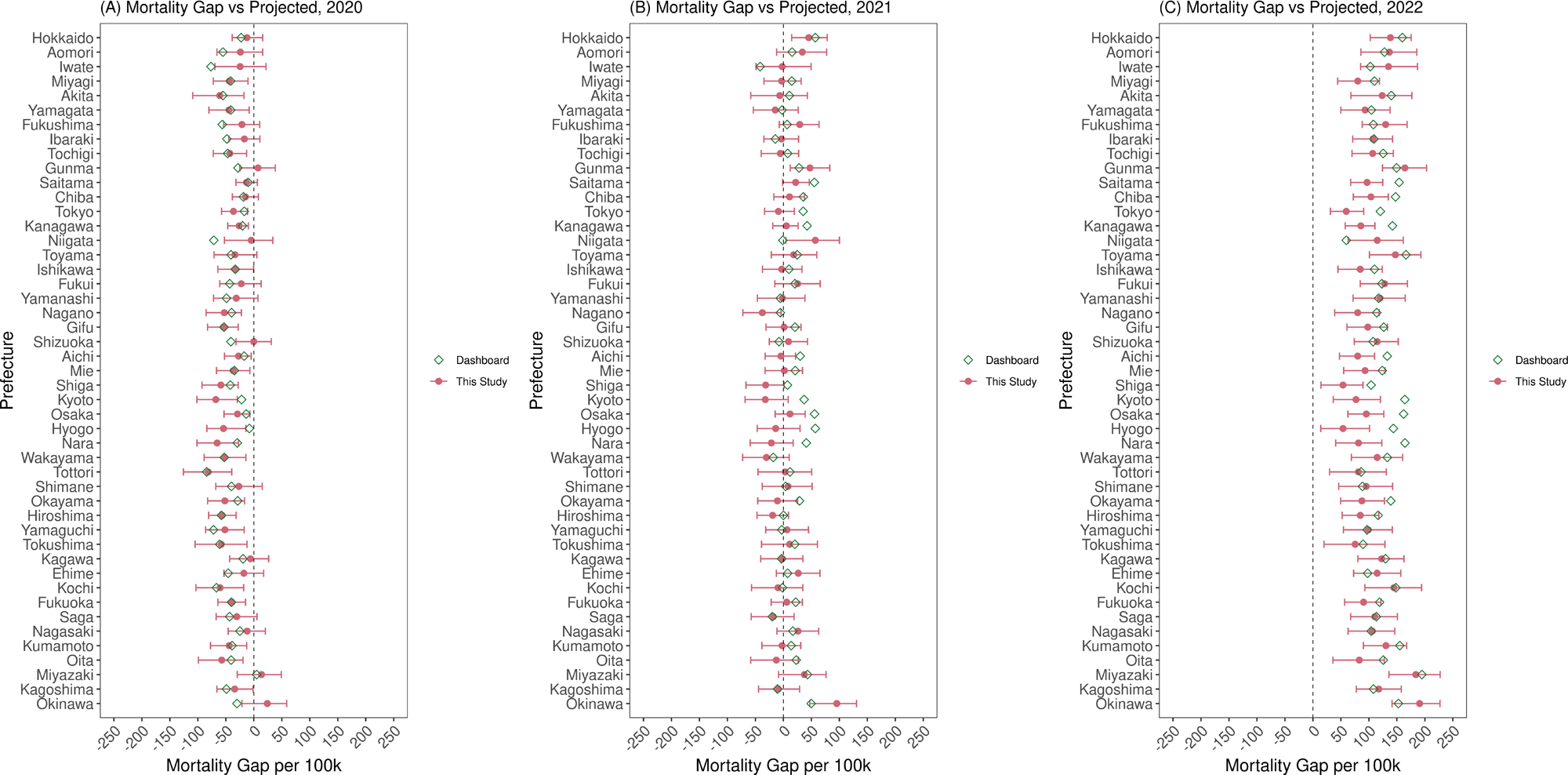
Gaps between observed and projected death counts per 100,000 population in 2020–2022. Gaps between observed and projected death counts per 100,000 population are shown in (A) 2020, (B) 2021, and (C) 2022 for all prefectures. In each panel and for each prefecture, red points and error bars show the median and 95% prediction interval, respectively. Green hollow diamonds represent the average yearly aggregated gap between observed and projected death counts provided in “Excess and Exiguous Deaths Dashboard in Japan.”^9^

Regarding gaps in life expectancy and death counts, results in the 5-year baseline scenario were not substantially different from the main scenario, as presented in the main text (see the Supplementary Figures 7-8).

## Discussion

In the present study, we proposed a framework to evaluate excess mortality by age and prefecture during the COVID-19 pandemic from 2020 to 2022 in Japan. By integrating demographic and statistical frameworks developed in previous studies, we projected the baseline mortality rates during 2000–2019 by extending the pre-pandemic trend up to 2022 and evaluated the difference between observed mortality rates and projected ones. Consideration for the age structures in mortality schedules in the present framework is a key distinction from established regression-based excess mortality estimations,^5,6,14–20^ making our framework an important addition to an array of conventional statistical and demographic approaches used in common excess mortality estimations. Although our analysis was limited to the mortality impact of the COVID-19 pandemic in Japan, the range of potential applications of our framework is wide because the proposed approach only requires yearly life table data that are readily available in many countries.

The key finding in our study was the substantial and steady increase in observed mortality rates and the loss of life expectancy at birth in 2021 and 2022, as illustrated in Figures 2–4. These results support our previous findings^31,32^ that mortality rates among individuals aged >30 years were significantly elevated during 2020–2022. Another important finding from our study was the importance of accounting for the underlying trend in the age-wise demographic structure, as suggested by the findings in Figure 5. Although our results basically align with the ExDeath Dashboard, substantially higher estimates are given in the ExDeath Dashboard for Tokyo, Osaka, and their neighboring prefectures, despite partial use of post-pandemic data. This finding might be due to our model accounting for baseline trends in age structure, in contrast to the Farrington algorithm used in the ExDeath Dashboard, which is designed to capture historical seasonal patterns. One future scope of methodological development might encompass the combination of age-structured models like ours with conventional statistical methodologies that also capture seasonality, which would be enabled when finer-scale chronological data become available.

Technically, the key contribution of our study is that we extended the framework by Alexander et al.,^24^ which combines matrix factorization of the mortality schedule matrix with time-varying regression analysis to enable excess mortality estimation. Although our primary goal was not to improve regional estimates as in Alexander et al., we projected future mortality schedules based on time-varying regression of the baseline period, which may likely reflect pre-pandemic demographic trends.

The present study has two limitations. First, our framework is designed to handle yearly life table data from Japan, which are usually reported with a time lag longer than a year; this naturally means that results cannot be obtained in a timely manner. However, our framework is also applicable with minor modifications when up-to-date data with shorter reporting intervals become available. Another important limitation is that we simply extended the pre-pandemic demographic trends to obtain the projected “baseline” mortality, which may have changed even in the absence of the COVID-19 pandemic. Even conventional regression-based approaches are not free of this limitation, but potential non-pandemic trend changes might have a larger impact on our approach because this method entirely omits post-pandemic data from the baseline estimation. However, our future projections of pre-pandemic mortality trends incorporated uncertainty, which should have alleviated such potential shortcomings by considering the cumulative uncertainty of the projections up to 2022. For further improvement regarding this issue, exploration of methodologies that are both practically feasible and ensure interpretability is warranted. Additional discussion regarding the length of the baseline period can be found in the Supplementary Discussion.

In conclusion, we successfully established a novel framework that combines demographic approaches with hierarchical Bayesian time-varying regression models to evaluate the mortality impact of the COVID-19 pandemic in Japan during 2020–2022. The application of this framework not only is limited to Japan or COVID-19 but also may serve as a tool for assessing excess mortality caused by unexpected external events, such as future pandemics, where longitudinal life table data are available.

## Acknowledgment

We thank Analisa Avila, MPH, ELS, of Edanz (https://jp.edanz.com/ac) for editing a draft of this manuscript.

## Funding Sources

Y.O. received funding from the SECOM Science and Technology Foundation. H.N. received funding from Health and Labour Sciences Research Grants (grant numbers 20CA2024, 21HB1002, 21HA2016, and 23HA2005), the Japan Agency for Medical Research and Development (grant numbers JP23fk0108612 and JP23fk0108685), JSPS KAKENHI (grant numbers 21H03198 and 22K19670), the Environment Research and Technology Development Fund (grant number JPMEERF20S11804) of the Environmental Restoration and Conservation Agency of Japan, the Daikin GAP Fund of Kyoto University, Japan Science and Technology Agency SICORP program (grant numbers JPMJSC20U3 and JPMJSC2105), the CREST program (grant number JPMJCR24Q3), and RISTEX program for Science, Technology, and Innovation Policy (grant number JPMJRS22B4). The funders had no role in the study design, data collection and analysis, decision to publish, or preparation of the manuscript.

## Conflict of interest

We declare that we have no conflicts of interest.

## Ethical approval statement

Ethical approval was not required because all data used in the present study did not include any personally identifiable information.

## Data availability

All data used in the present study are publicly available from the Japanese Mortality Database (https://www.ipss.go.jp/p-toukei/JMD/index-en.asp) and from the “Excess and Exiguous Deaths Dashboard in Japan” (URL: https://exdeaths-japan.org/en/). The supplementary files include the summary data from the results of our analyses.

## Notes

### Competing Interest Statement

The authors have declared no competing interest.

