## supplementary material for "Demographic analysis of mortality changes associated with the COVID-19 pandemic in Japan, 2020–2022"

**Supplementary Materials**

**Supplementary Methods 1: Description of statistical methods used in the present study.**

The age-stratified mortality schedules for 2000–2019 in Japan were used to create a mortality schedule matrix. We let the log-transformed form of this matrix be **M** with 20 rows and 22 columns (corresponding to the number of years and age groups, respectively). Singular value decomposition of **M** is in the form

$$\mathbf{M} = \mathbf{U\Sigma V}^{\mathrm{T}}, \qquad \text{(S1)}$$

where **U** is a 20 by 20 matrix representing a time-dependent component, $\mathbf{\Sigma}$ is a 20 by 22 matrix generating singular values, and **V** is a 22 by 22 matrix describing the age structure, respectively. Note that $\mathbf{\Sigma}_{p,p}$, the (p, p)-th element of $\mathbf{\Sigma}$, is the p-th singular value of **M**. The p-th column of **V**, $Y_x^{(p)}$, represents the age–mortality structure corresponding to the p-th singular value $\mathbf{\Sigma}_{p,p}$.

**Hierarchical time-varying regression model**

The mortality rate in age group *x* and in prefecture *a* in year *t* was modeled as a linear combination of $Y_x^{(1)}$, $Y_x^{(2)}$, and $Y_x^{(3)}$:

$$\log\left(m_{x,a,t}\right) = \sum_{p=1}^{3} \beta_{a,t}^{(p)} Y_x^{(p)} + u_{x,a}, \qquad (2)$$

where $\beta_{a,t}^{(p)}\ (p = 1, 2, 3)$ are the coefficients in year *t*. The term $u_{x,a}$ in equation (2) is the random effects by prefecture in age *a*, which were assumed to be time-invariant. For $\beta_{a,t}^{(p)}$ and $u_{x,a}$, we assumed

$$\psi_{\beta|a,t}^{(p)} = \beta_{a,t}^{(p)} - \overline{\beta_t^{(p)}} \sim Normal\left(0, \sigma_{\psi_\beta,t}^{(p)}\right), \qquad (3)$$

$$u_{x,a} \sim Normal(0, \sigma_x), \qquad (4)$$

where $\beta_{a,t}^{(p)}$ is modeled as the sum of the national averages $\overline{\beta_t^{(p)}}$ and random effects $\psi_{\beta|a,t}^{(p)}$ in year *t*. Random effects $u_{x,a}$ are assumed to be time-invariant, and $u_{x,a}$ in each age group is assumed to be centered around zero with uncertainty represented by $\sigma_x$.

Regarding the above-mentioned $\overline{\beta_t^{(p)}}$, the following likelihoods were also considered given that $\overline{\beta_t^{(p)}}$ should in principle closely align with $\boldsymbol{\Sigma}_{p,p}\mathbf{U}_{t,p}$, i.e., the product of the p-th singular value of **M** and the (t, p)-th element of **U**:

$$\overline{\beta_t^{(p)}} \sim Normal\left(\boldsymbol{\Sigma}_{p,p}\mathbf{U}_{t,p}, \sigma_{\overline{\beta}}^{(p)}\right), \tag{5}$$

where $\sigma_{\overline{\beta}}^{(p)}$ represents parameters that correspond to standard deviations and are therefore to be estimated.

To account for the mortality impact of the Tohoku earthquake on those aged 0–4 years in 2011, we introduced the parameters $\epsilon_{x,a}$ that are only defined for age groups 0 and 1–4 years of 2011 in Iwate, Miyagi, and Fukushima. Using $\epsilon_{x,a}$, the overall mortality $m'_{x,a,t}$ is

$$m'_{x,a,t} = m_{x,a,t} + \epsilon_{x,a}$$
$$when\ t = 2011, x \in \{0, 1\text{-}4\}, a \in \{Iwate, Miyagi, Fukushima\}, \tag{6}$$

and

$$m'_{x,a,t} = m_{x,a,t} : otherwise. \tag{7}$$

**Statistical inference of temporal changes in mortality**

We let $D_{x,a,t}$ and $P_{x,a,t}$ be the death counts and exposure-to-risk population in age group *x*, prefecture *a*, in year *t*. Using $m'_{x,a,t}$, the likelihoods of observing $D_{x,a,t}$ can be described as:

$$D_{x,a,t} \sim Poisson\left(m'_{x,a,t} P_{x,a,t}\right). \tag{8}$$

By assuming independence of the observation process of death counts in age groups, the following likelihood was also considered:

$$\sum_x D_{x,\,a,\,t} \sim Poisson\left(\sum_x m'_{x,\,a,\,t} P_{x,\,a,\,t}\right). \tag{9}$$

Regarding deaths documented as directly associated with the 2011 earthquake, the likelihood of observing death counts in those aged 0–4 years owing to the earthquake was modeled as follows:

$$D^{EQ}_{0-4,\,a,\,2011} \sim Poisson\big(\epsilon_{0,a} P_{0,\,a,\,2011} + \epsilon_{1-4,a} P_{1-4,\,a,\,2011}\big), \quad a \in \{Iwate, Miyagi, Fukushima\}. \tag{10}$$

The statistical model for the temporal changes in $\overline{\beta^{(p)}_t}$ was assumed as a second-order random walk:

$$\overline{\beta^{(p)}_{t+1}} - \overline{\beta^{(p)}_t} \sim Normal\left(\overline{\beta^{(p)}_t} - \overline{\beta^{(p)}_{t-1}}, \sigma^{(p)}_{\beta,time}\right) \tag{11}$$

The above-mentioned likelihoods were used in the Markov chain Monte Carlo conducted to estimate parameters $\overline{\beta^{(p)}_t}, \sigma^{(p)}_{\psi_\beta,t}, u_{x,\,a}$, $\sigma^{(p)}_{\beta}$, $\sigma^{(p)}_{\overline{\beta}}, \sigma_x, \epsilon_{x,a}$, and $\sigma^{(p)}_{\beta,time}$. The prior distributions used are described in Supplementary Methods 2. For each of the four chains run, we generated 500 warm-up samples followed by 1,000 posterior samples. To confirm that the chain converged, we ensured that the R-hat value was below 1.01.

**Future projection of pre-pandemic mortality trends**

For projection of $\beta^{(p)}_{a,\,t}$ during 2020–2022, we first projected $\overline{\beta^{(p)}_t}$ as follows. Assuming that the change rate of $\overline{\beta^{(p)}_t}$ from 2018 to 2019 was maintained at $\Delta^{(p)}_{\overline{\beta}}$:

$$\Delta^{(p)}_{\overline{\beta}} = \overline{\beta^{(p)}_{2019}} - \overline{\beta^{(p)}_{2018}}, \tag{12}$$

we obtain

$$\overline{\beta^{(p),projected}_{2022}} = \overline{\beta^{(p)}_{2019}} + 3\Delta^{(p)}_{\overline{\beta}}, \tag{13}$$

$$\overline{\beta_{2021}^{(p),projected}} = \overline{\beta_{2019}^{(p)}} + 2\Delta_{\overline{\beta}}^{(p)},$$

$$\overline{\beta_{2020}^{(p),projected}} = \overline{\beta_{2019}^{(p)}} + \Delta_{\overline{\beta}}^{(p)}.$$

For the projection of $\psi_{\beta|a,2022}^{(p)}$, $\psi_{\beta|a,2021}^{(p)}$, and $\psi_{\beta|a,2020}^{(p)}$, we set these to the same value as $\psi_{\beta|a,2019}^{(p)}$. By conducting the same procedure for all prefectures, we obtained projected mortality rates at the national level, $\overline{m_{x,\,t}^{\prime,projected}}$, and at prefectural levels, $m_{x,\,a,\,t}^{\prime,projected}$, up to 2022.

It remained unclear how long the baseline period should be. As a sensitivity analysis, a “5-year baseline scenario” was tested, where 5-year-averages of the changes in $\overline{\beta_t^{(p)}}$ and the values of $\psi_{\beta|a,t}^{(p)}$ by prefecture were used for projection beyond 2020. The results can be found in Supplementary Figures 4-8.

**Evaluation of the post-pandemic mortality impact**

Predicted distributions of $D_{x,\,a,\,t}^{projected}$ were obtained by random sampling from the following Poisson distributions:

$$D_{x,\,a,\,t}^{projected} \sim Poisson\left(m_{x,\,a,\,t}^{\prime,projected} P_{x,\,a,\,t}\right):\ t = 2020 - 2022, \tag{14}$$

and the predicted values of $m_{x,\,a,\,t}^{\prime,projected}$ were calculated as $D_{x,\,a,\,t}^{projected}/P_{x,\,a,\,t}$, accordingly. By comparing $D_{x,\,a,\,t}^{projected}$ to the observed death counts $D_{x,\,a,\,t}$, the gaps between observed and projected “pre-pandemic” baseline death counts were calculated as:

$$\Delta D_{x,a,t} = D_{x,\,a,\,t} - D_{x,\,a,\,t}^{projected}:\ t = 2020 - 2022, \tag{15}$$

which can be interpreted as excess or exiguous deaths.

The gaps between observed and projected life expectancy at birth, $\Delta e_{0|a,t} = e_{0|a,t}^{observed} - e_{0|a,t}^{projected}$, were also evaluated by calculating $e_{0|a,t}^{projected}$ using predicted values of $m_{x,\,a,\,t}^{\prime,projected}$ $(t = 2020\text{-}2022)$. On the basis of these mortality rates, projected life expectancy was calculated for each prefecture.

**Supplementary Methods 2: Priors used for Bayesian inference by Markov chain Monte Carlo.**

For $\sigma_{\overline{\beta}}^{(p)}, \sigma_{\beta}^{(p)}, \sigma_x$, we used:

$$\sigma_{\overline{\beta}}^{(p)}, \sigma_{\beta}^{(p)}, \sigma_x \sim Inverse\ Gamma(2, 2), \tag{S1}$$

and for $\sigma_{\beta,time}^{(p)}$, we used:

$$\sigma_{\beta,time}^{(p)} \sim Half\ Cauchy(0, 1) \tag{S1}$$

For $\epsilon_{x,a}$, we used informative priors based on the observed mortality change from 2010 to 2011 in Iwate, Miyagi, and Fukushima prefectures:

$$\begin{gathered} \mathrm{Log}\ (\epsilon_{0,a}) \sim Normal\left(\log\left(m_{0,\,a,\,2011}^{observed}\right) - \log\left(m_{0,\,a,\,2010}^{observed}\right), 0.1\right), \\ \log\left(\epsilon_{1-4,a}\right) \sim Normal\left(\log\left(m_{1-4,\,a,\,2011}^{observed}\right) - \log\left(m_{1-4,\,a,\,2010}^{observed}\right), 0.1\right) \\ a \in \{Iwate, Miyagi, Fukushima\} \end{gathered} \tag{S2}$$

**Supplementary Methods 3: Reason for choosing three leading singular values (and corresponding right singular vectors).**

Choosing three leading singular values is in line with Alexander et al. Those authors stated the merit of choosing three as allowing flexibility in mortality schedules.

In addition to this, and with the aim to enhance comparability and interpretability in the context of the previous study, there are three reasons for choosing three leading singular values:

1) The three right singular vectors represent demographically meaningful patterns. Namely, the first represents the baseline mortality level across ages, the second represents high mortality in children and elderly adults, which may require higher levels of health care to be averted, and the third may reflect high child/adolescent mortality owing to causes such as accidents or suicide.

2) Choosing three leading vectors facilitates nearly an exact approximation of the original mortality schedule matrix. When considering $\mathbf{M}_{(3)} = \mathbf{U}\boldsymbol{\Sigma}_{(\mathbf{3})}\mathbf{V}^{\mathrm{T}}$, the approximation of $\mathrm{M}$ by up to three leading singular values, where

$$\boldsymbol{\Sigma}_{(\mathbf{3})|\boldsymbol{i},\boldsymbol{j}} = \begin{cases} \boldsymbol{\Sigma}_{p,p} : \text{ if } i = j = p \ \leq 3, \\ \quad 0 : \text{ otherwise,} \end{cases}$$

the Frobenius norms of $\mathbf{M}_{(3)}$ and $\mathbf{M}$ satisfies

$$\frac{\left\|\mathbf{M}_{(3)}\right\|_F^2}{\|\mathbf{M}\|_F^2} > 0.9999,$$

which means an almost exact match between these two matrices.

3) Differences between neighboring singular values in log-scale plateaued after differences in the 3rd to 4th singular values (Supplementary Figure 1).

**Supplementary Discussion**

How to choose the length of the baseline period is an open question, and this length has been arbitrarily chosen in conventional approaches to excess mortality. The present study also involved the issue of arbitrary choice of this length; therefore, we conducted additional analysis using the projections based on averages of prefectural random effects and yearly changes in national-level coefficients over 5 years.

As presented in Supplementary Figures 4–8, we confirmed that projections using the 5-year baseline period did not affect our result regarding the mortality impact of COVID-19 that were evaluated as gaps in mortality rates, life expectancy, or absolute death counts. This finding supports the robustness of our approach regarding the length of the baseline period.

**Supplementary Figure 1. Singular values of M and its log-difference.**

Panel (A) shows the singular values of M in log scale. Panel (B) shows the gap between neighboring log-scaled singular values.

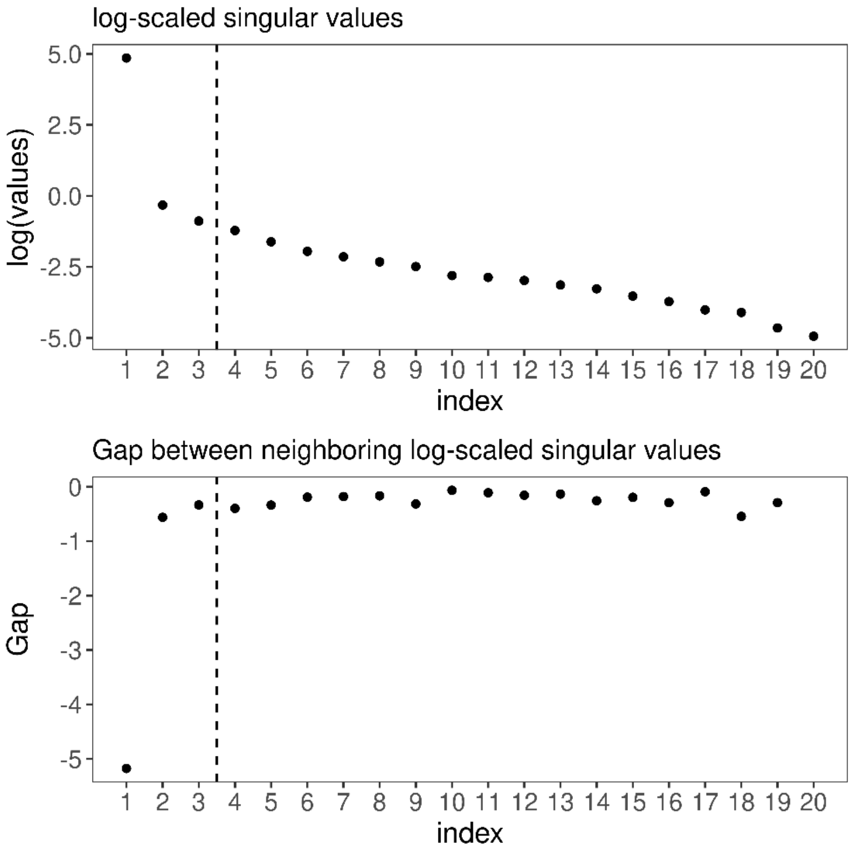

**Supplementary Figure 2. Three right singular vectors used in the model.** Three right singular vectors representing (A) the baseline mortality, (B) higher infant and adult mortality, and (C) higher child and adolescent mortality are visualized.

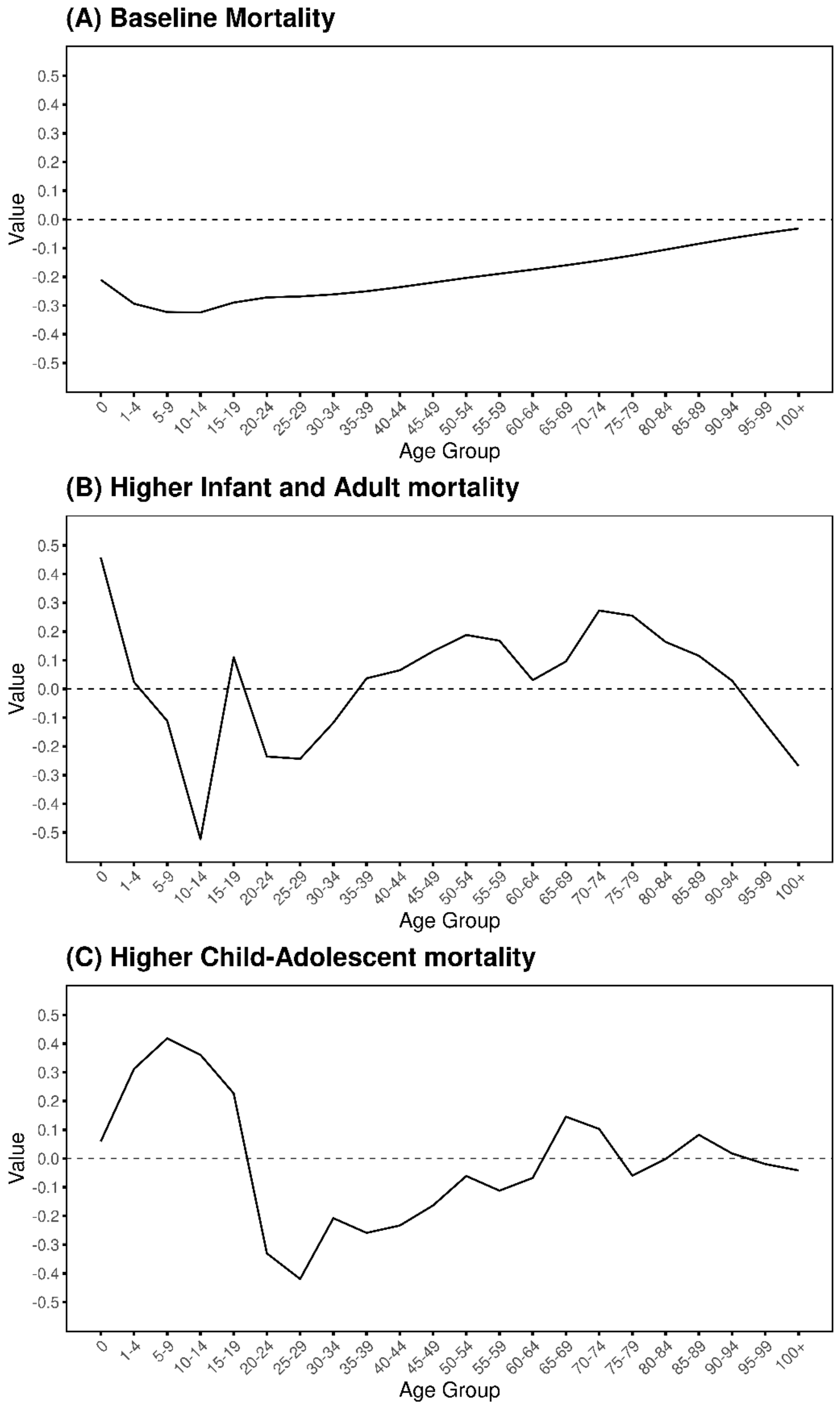

**Supplementary Figure 3. Coefficient of the three leading vectors $\boldsymbol{Y_x^{(1)}}$, $\boldsymbol{Y_x^{(2)}}$, and $\boldsymbol{Y_x^{(3)}}$.** Coefficients of the three right singular vectors representing (A) the baseline mortality, (B) higher infant and adult mortality, and (C) higher child and adolescent mortality are visualized together with the projected values beyond 2020.

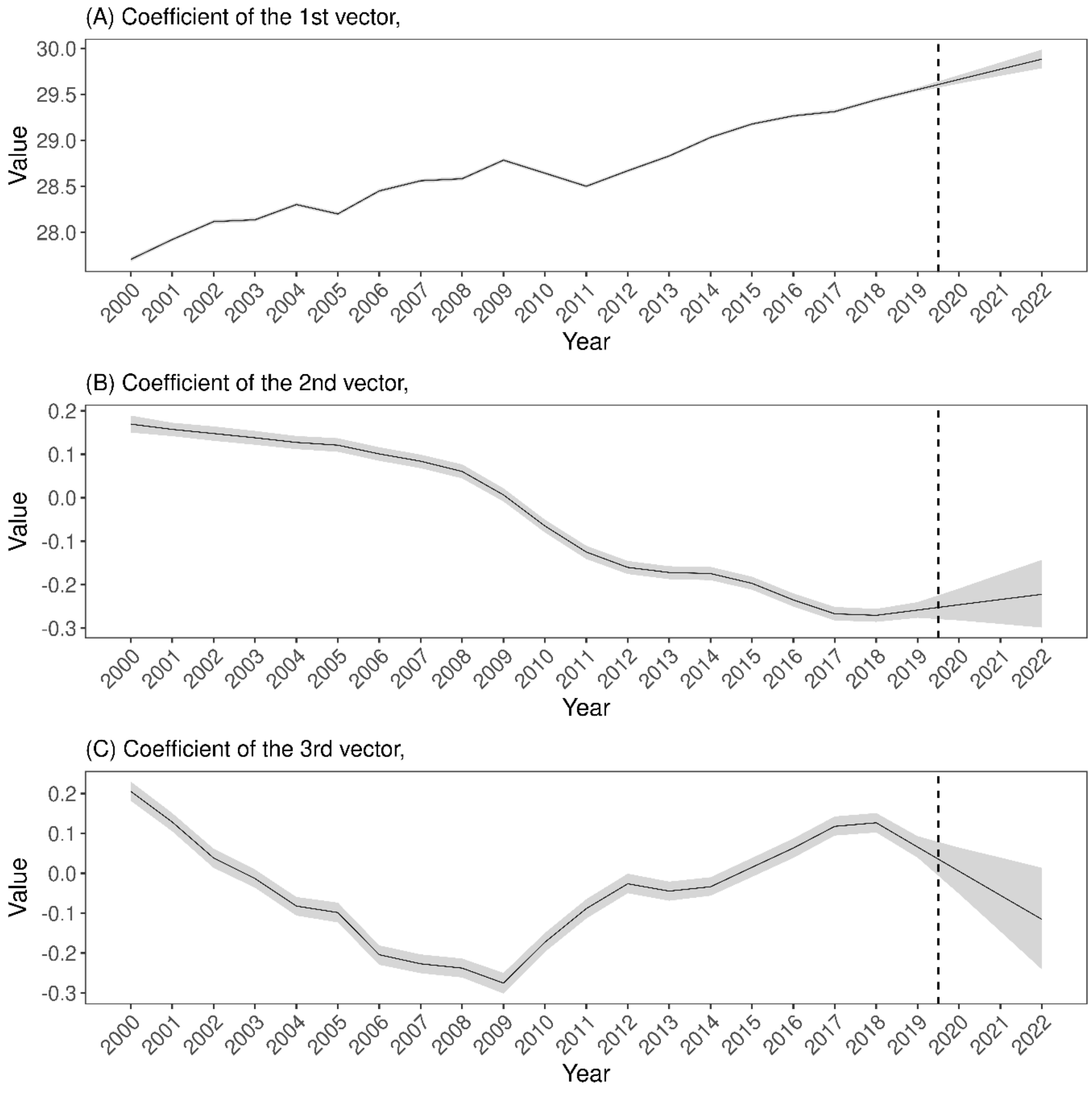

**Supplementary Figure 4. Coefficient of the three leading vectors $Y_x^{(1)}$, $Y_x^{(2)}$, and $Y_x^{(3)}$ in the 5-year baseline scenario.**

This figure is essentially the same as Supplementary Figure 3, except that the projections in each panel beyond 2020 are based on average changes during 2014–2019.

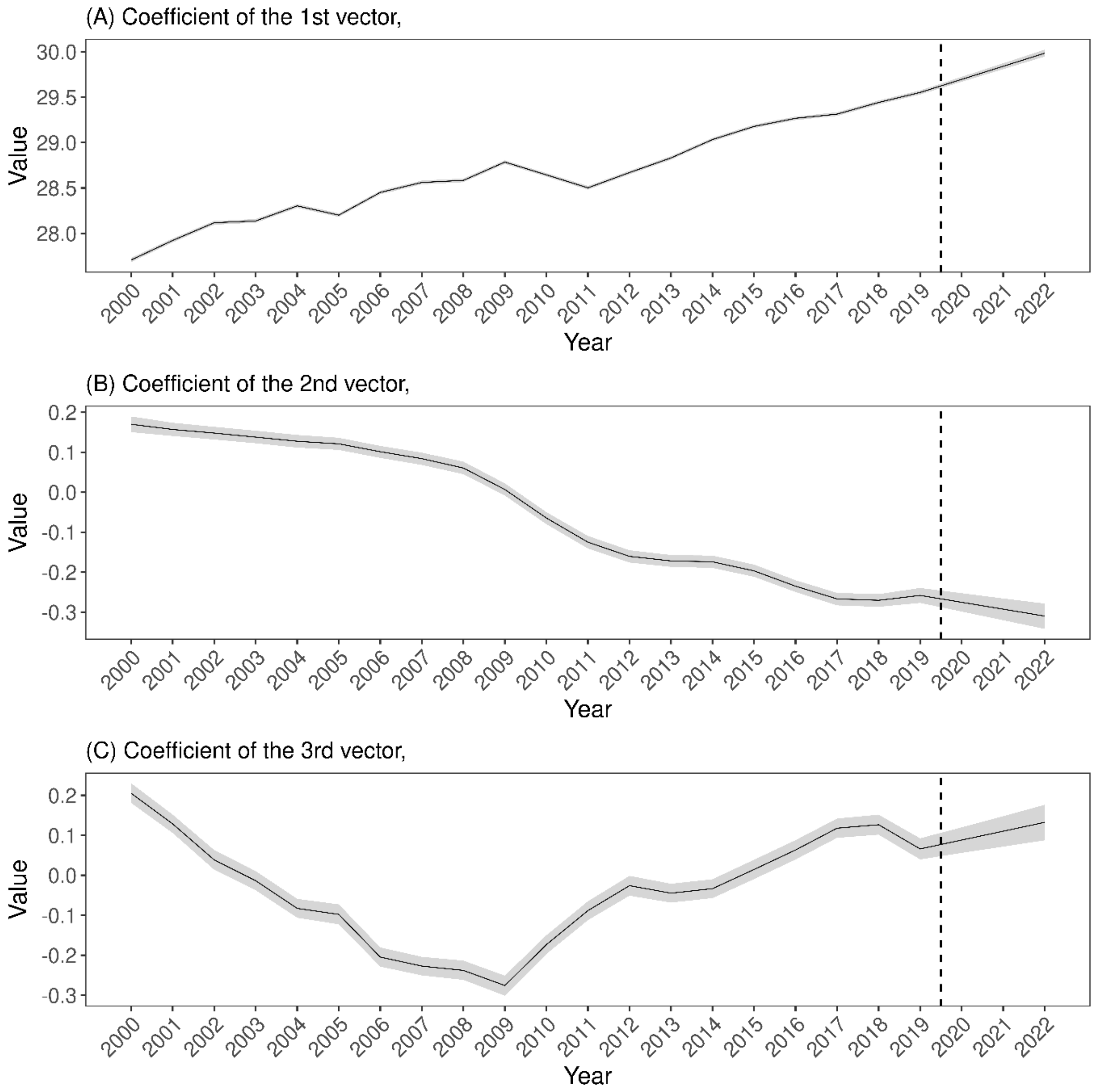

**Supplementary Figure 5. Gaps between observed and projected mortality rates by age group during 2020–2022 in the 5-year baseline scenario.**

The red points with error bars show the median and 95% prediction intervals of the gaps between observed and projected (log-scaled) mortality rates in (A) 2020, (B) 2021, and (C) 2022.

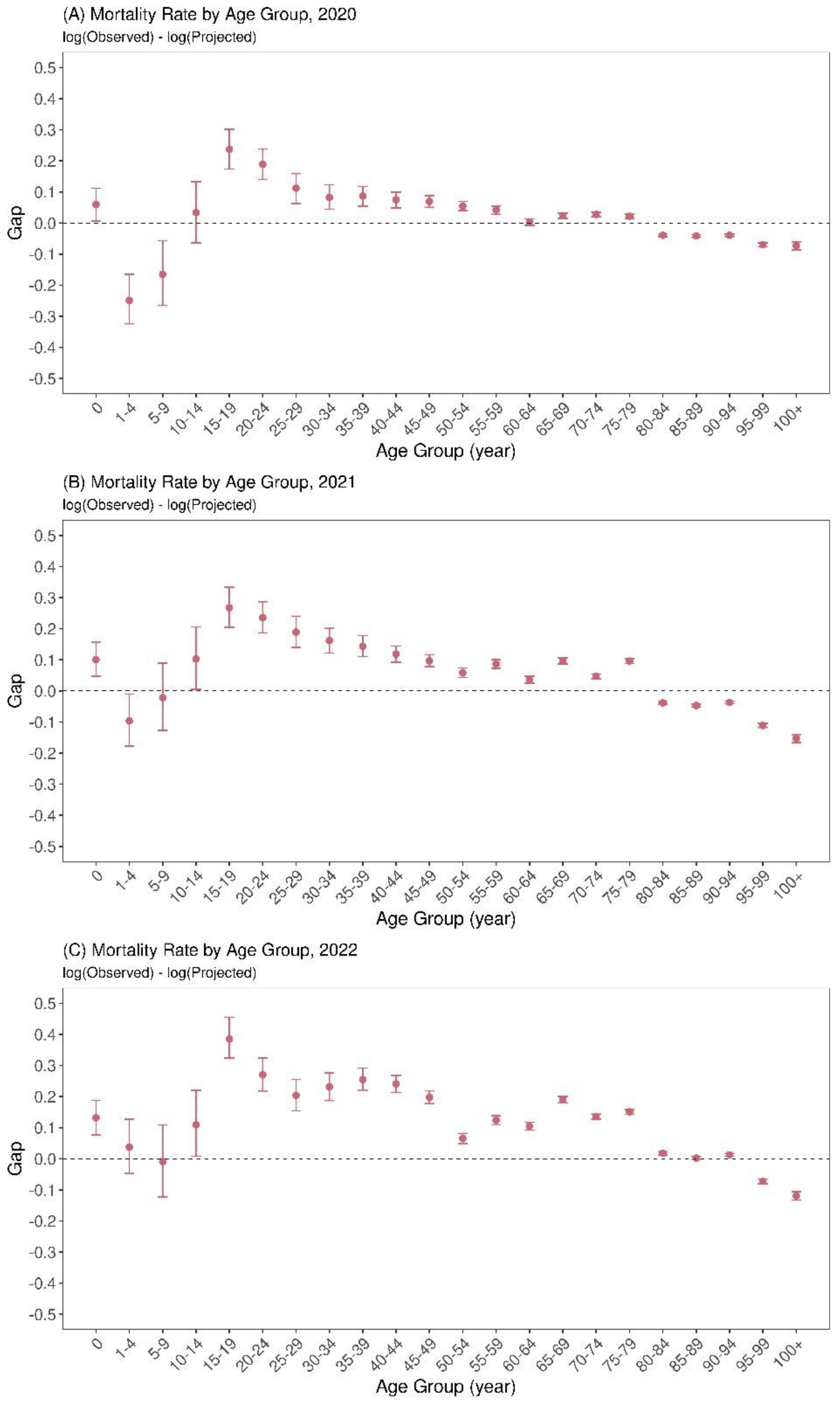

**Supplementary Figure 6. Heatmaps of the gap between observed and projected mortality rates by prefecture during 2020–2022 in the 5-year baseline scenario.** Cells represent gaps between the observed and the median of projected (log-scaled) mortality rates in (A) 2020, (B) 2021, and (C) 2022 for all prefectures. Here, the projections are based on the “5-year baseline scenario” in the main text. Cells with colors closer to dark blue show lower than expected (projected) mortality rates whereas those closer to solid red indicate higher than expected mortality rates.

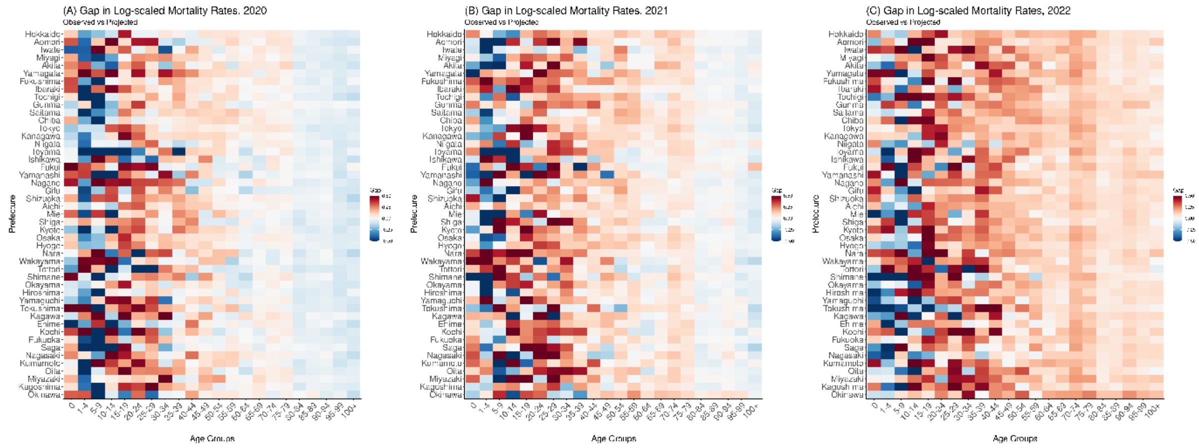

**Supplementary Figure 7. Gaps between life expectancy at birth calculated from observed and projected mortality rates during 2020–2022 in the 5-year baseline scenario.**

Gaps between the observed and projected life expectancy at birth are shown in (A) 2020, (B) 2021, and (C) 2022 for all prefectures. Here, the projections are based on the "5-year baseline scenario" in the main text. In each panel and for each prefecture, red points and error bars show the median and 95% prediction interval, respectively.

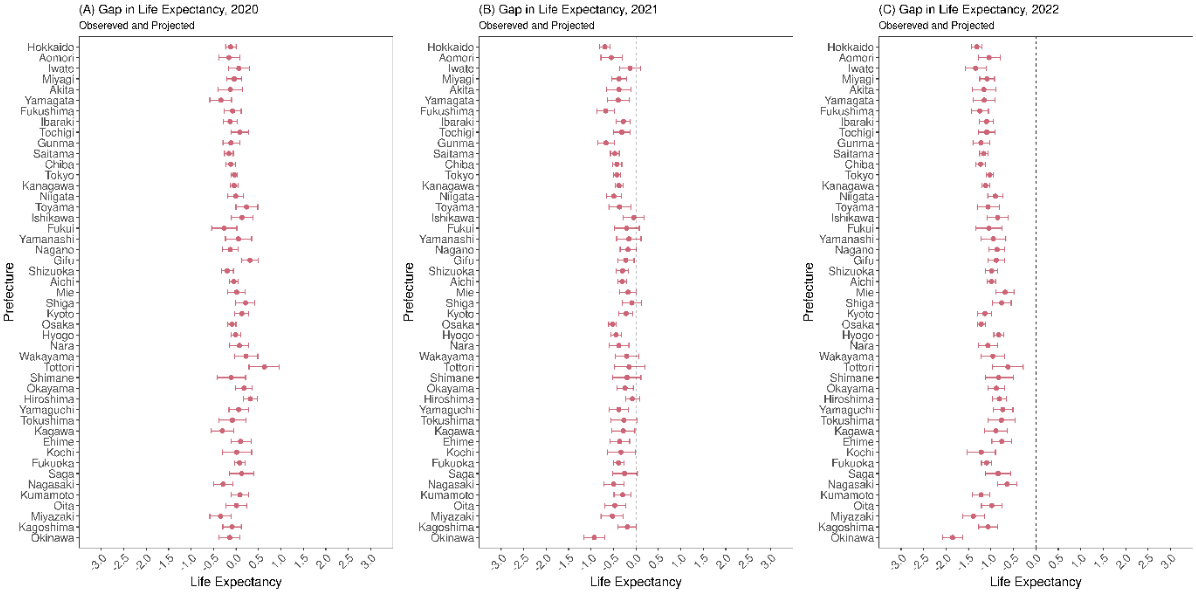

**Supplementary Figure 8. Gaps between observed and projected death counts per 100,000 population during 2020–2022 in the 5-year baseline scenario.**

Gaps between the observed and projected death counts per 100,000 population are shown in (A) 2020, (B) 2021, and (C) 2022 for all prefectures. Here, the projections are based on the "5-year baseline scenario" in the main text. In each panel and for each prefecture, red points and error bars show the median and 95% prediction interval, respectively. Green hollow diamonds represent the average yearly aggregated gap between observed and projected death counts provided by the "Excess and Exiguous Deaths Dashboard in Japan".

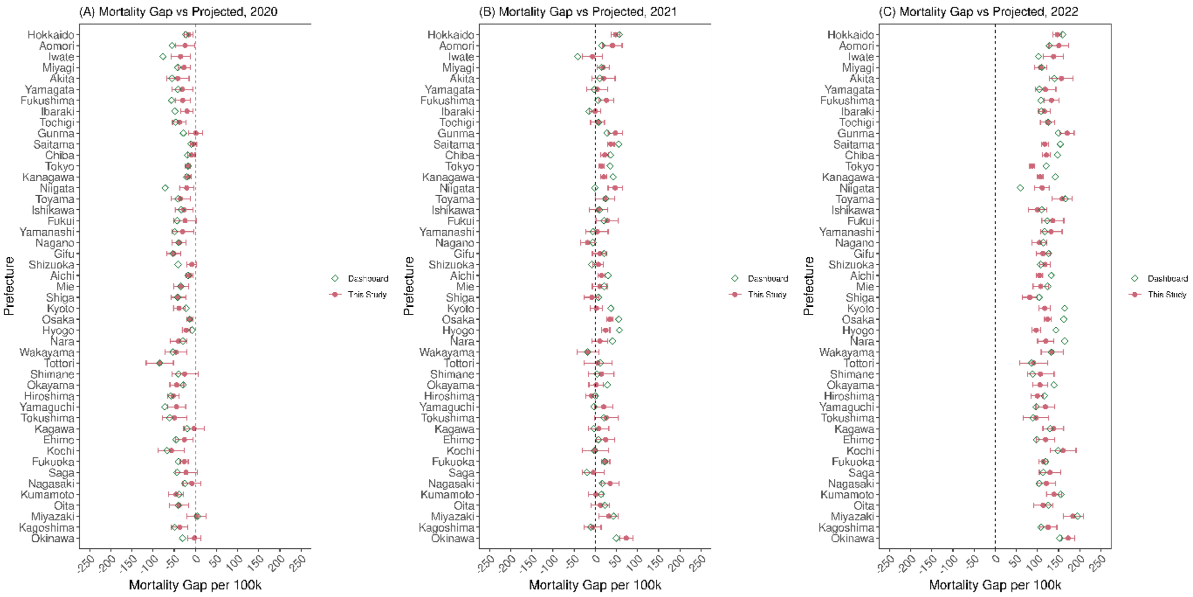

**Supplementary Table. Posterior estimates of $\boldsymbol{\sigma^{(p)}_{\beta,time}}$ and $\boldsymbol{\epsilon_{x,a}}$.**

| Parameters | Estimated values (95% CrI*) |
|---|---|
| $\sigma^{(1)}_{\beta,time}$ | 0.182 (0.133, 0.266) |
| $\sigma^{(2)}_{\beta,time}$ | 0.022 (0.014, 0.037) |
| $\sigma^{(3)}_{\beta,time}$ | 0.064 (0.042, 0.102) |
| $\epsilon_{0,Iwate}$ | 0.025 (0.022, 0.026) |
| $\epsilon_{1-4,Iwate}$ | 0.008 (0.007, 0.009) |
| $\epsilon_{0,Miyagi}$ | 0.015 (0.014, 0.017) |
| $\epsilon_{1-4,Miyagi}$ | 0.005 (0.005, 0.006) |
| $\epsilon_{0,Fukushima}$ | 0.015 (0.014, 0.016) |
| $\epsilon_{1-4,Fukushima}$ | 0.005 (0.004, 0.005) |

*CrI, Credible Interval.

**Supplementary Data 1. Estimates of the prefectural random effects in log-scaled mortality rates by age group.**

**Supplementary Data 2. Estimates of $\psi_{\beta|a,t}^{(p)}$, the prefectural random effects around $\overline{\beta_t^{(p)}}$.**

**Supplementary Data 3. Estimates of $\sigma_{\psi_\beta,t}^{(p)}$, the standard deviation of $\psi_{\beta|a,t}^{(p)}$.**

**Supplementary Data 4. Projected life expectancy at birth by prefecture during 2020–2022.**

**Supplementary Data 5. Projected gaps between observed and projected deaths per 100,000 population by prefecture during 2020–2022.**

**Supplementary Data 6. Projected life expectancy at birth by prefecture during 2020–2022 in the 5-year baseline scenario.**

**Supplementary Data 7. Projected gaps between observed and projected deaths per 100,000 population by prefecture during 2020–2022 in the 5-year baseline scenario.**
